# A new incidence peak of childhood narcolepsy type 1 in 2013: a new perspective on the role of influenza virus?

**DOI:** 10.1101/2019.12.31.19016246

**Authors:** Zhongxing Zhang, Jari Gool, Rolf Fronczek, Claudio L. Bassetti, Yves Dauvilliers, Geert Mayer, Giuseppe Plazzi, Fabio Pizza, Joan Santamaria, Markku Partinen, Sebastiaan Overeem, Rosa Peraita-Adrados, Antonio Martins da Silva, Karel Sonka, Rafael del Rio-Villegas, Raphael Heinzer, Aleksandra Wierzbicka, Peter Young, Birgit Högl, Mauro Manconi, Eva Feketeova, Johannes Mathis, Teresa Paiva, Francesca Canellas, Michel Lecendreux, Christian R. Baumann, Lucie Barateau, Carole Pesenti, Elena Antelmi, Carles Gaig, Alex Iranzo, Laura Lillo-Triguero, Pablo Medrano-Martínez, José Haba-Rubio, Corina Gorban, Gert-Jan Lammers, Ramin Khatami, on behalf of the EUROPEAN NARCOLEPSY NETWORK (EU-NN)

## Abstract

Increased incidence rates of narcolepsy type 1 (NT1) after the 2009-2010 H1N1 influenza pandemic (pH1N1) have been reported world-wide. While some European countries found an association between the NT1 increase and H1N1 vaccination Pandemrix, reports from Asian countries suggested the H1N1 virus rather than Pandemrix to be linked with the increase of new NT1 cases. We analyzed the number of de-novo NT1 cases in the last two decades until 2016 using the European Narcolepsy Network (EU-NN) database. Using robust data-driven modelling approaches we confirmed the peak of NT1 incidence in 2009-2010 pH1N1 and identified a new peak in 2013 that is age-specific for children/adolescents. Most of these de-novo cases showed a subacute disease onset consistent with an immune-mediated type of narcolepsy, which is most likely not related to Pandemrix vaccination that was used in 2009-2010, but may have been triggered by some new epidemiological event in Europe. Our finding of an unexpected peak in de-novo children narcolepsy in 2013 provides a unique opportunity to develop new hypotheses, such as considering other (influenza) viruses to further investigate the pathophysiology of immune-mediated narcolepsy.

## Introduction

Narcolepsy type 1 (NT1) is a rare disabling brain disorder (prevalence of 0.02-0.05%^1, 2^). It is characterized by the presence of excessive daytime sleepiness (EDS) and cataplexy, and/or a selective loss or dysfunction of orexin neurons. NT1 may arise from the complex interactions of genetic and environmental factors that trigger an immune-mediated response targeting orexin neurons in the lateral hypothalamus^3^. The increased incidence rates (IRs) of NT1 after Pandemrix (GlaxoSmithKline Biologicals, Wavre, Belgium), an AS03-adjuvanted monovalent pandemic H1N1 influenza (pH1N1) vaccine, has been repeatedly reported in several European countries including Finland, Sweden, France, England, Ireland and Norway^4-7^ after the 2009-2010 pH1N1, leading to a suspicion of the association between Pandemrix and the development of narcolepsy. However, the increased IRs of narcolepsy have also been previously reported in East Asia region not widely using Pandemrix such as in mainland China^8, 9^ and Taiwan^10^. In South Korea scientists found no association between the non-adjuvanted and MF59-adjuvanted H1N1 vaccines and narcolepsy at all ^11, 12^, and the IR was actually highest in pre-pandemic period^11^. Thus, the pathophysiological links between narcolepsy and pH1N1 vaccination or the virus itself as potential environmental factors, are still not completely understood even 10 years after the pH1N1 influenza in 2009.

One reason for the unclear association between narcolepsy and exposure to the vaccine or the virus is that we are still lacking data collected in the years following the pH1N1 pandemic. Narcolepsy is a clinical syndrome with either severe and abrupt symptom onset or a more progressive development with several months or years between daytime sleepiness and cataplexy onset. People presenting EDS in 2009-2010 could be diagnosed with narcolepsy a few years later due to the progressive development of syndromes or lack of awareness of the diseases^13^. The delayed diagnosis can cause a bias when investigating the temporal association between vaccination/pH1N1 virus and narcolepsy, and help to explain false results in previous studies^6^. Therefore, the data of NT1 onset in the years after 2010 will contribute to clarify the confounding between the influenza virus itself and the vaccination in inducing narcolepsy. Moreover, if increased NT1 incidence during influenza seasons after 2010 would be identified, it could indicate that an influenza virus, some other agent or a combination of different immunological triggers (e.g. a viral infection combined with a streptococcal infection) may also be associated with an onset of NT1, in addition to the Pandemrix^6^. European Medicines Agency recommended restricting use of Pandemrix but the H1N1 virus has still circulated after 2009-2010 pH1N1. Currently only limited data after 2010 (i.e., only until 2012 or 2013) were available from some countries ^9, 14, 15^. In a report from the Finnish Department of Health and Welfare the incidence started to decrease significantly after the second year since Pandemrix vaccination^16^. This means that a possible later increase in incidence (after end of 2011) may be due to H1N1 influenza virus or some other agent that have been used after 2011.

The European narcolepsy network (EU-NN) is an association of leading European sleep centers. It launched the first prospective European web-based database for narcolepsy and related disorders which allows collection, storage and dissemination of data on narcolepsy in a comprehensive and systematic way^13^. The current EU-NN database includes data of 994 NT1 patients from 26 sleep centers diagnosed from 1980s to 2018. In this study, we use a data-driven approach to compare the numbers of NT1 patients presenting symptoms before, during and a few years post the 2009-2010 pH1N1 in EU-NN database. Giving that the EU-NN database is of good quality control^13, 17^, i.e., the structure and data acquisition is standardized according to EU-NN regulations and the data are collected from most of the leading European sleep centers guaranteeing reliable diagnosis of NT1, we believe that this study can deepen our knowledge of the etiology of pH1N1 or vaccination associated narcolepsy in 2009-2010. Using the EU-NN database we aim to 1) test whether we could replicate the Chinese findings of increased NT1 including those cases that haven‘t been vaccinated with Pandemrix in 2009-2010 in a pan-European database; 2) test whether the increased NT1 peak was unique for the 2009-2010 influenza season or repeated over time, compatible with the so called multiple hit hypothesis. Recurrent increased IRs would indicate that immune-mediated narcolepsy is not necessarily specific to H1N1 virus or Pandemrix; 3) identify possible differences between different age groups related to the increased NT1 peak.

## Methods

Each center of EU-NN has obtained ethical approval for publishing the patients’ data for scientific purpose by a national Institutional Review Board before entering patients (please refer to the section ‘Data access policy, ethics and security specifications’ of reference^13^). The scientific review committee of EU-NN has approved the study protocol. All methods are in accordance with the relevant guidelines and regulations. All patients have provided their informed consent to be entered into the EU-NN database and their data can be used for scientific studies.

Patients with NT1 were diagnosed according to the third edition of International Classification of Sleep Disorders (ICSD-3). Patients with daily periods of the irrepressible need to sleep or daytime lapses into sleep occurring for at least 3 months and the presence of at least one of the following:

1. Cataplexy and mean sleep latency of 8 minutes or less and at least 2 sleep onset rapid eye movement periods (SOREMPs) on a multiple sleep latency test (MSLT). A SOREMP on the preceding nocturnal polysomnography may replace one of the SOREMPs on the MSLT.
2. Cerebrospinal fluid (CSF) hypocretin-1 concentration, measured by immunoreactivity, is after conversion to Stanford values either 110 pg/mL or less, or less than one third of mean values obtained in normal subjects using the same standardized assay.

The following criteria were used exclude NT1 patients from the EU-NN database for our analysis:

1. Patients with an indefinite diagnosis. The EU-NN database contains a variable on certainty of clinical diagnosis. The clinicians were asked rate their diagnosis certainty on a 3-level basis (probable, possible and definite). All 148 patients with a possible or probably NT1 diagnosis certainty were excluded.
2. The 33 patients with missing information of the year of starting EDS were excluded.
2. The 250 patients with onset of EDS before 1995 were excluded.
3. Only countries with more than 30 patients were selected. Patients from Austria (n=13), Poland (n=14), Portugal (n=18), Scotland (n=1), Slovakia (n=9) were excluded due to relative smaller number of patients in these countries.

In total, 508 patients (f:230, m:278, mean age of starting EDS: 22.01±12.79 years) from Czech Republic (n=31), Finland (n=42), France (n=114), Germany (n=84), Italy (n=90), Netherlands (n=58), Spain (n=53) and Switzerland (n=36) were included. The latest onset of EDS in these patients was in 2016.

We chose the year of starting EDS as the disease onset year, as EDS in general was the first developed symptom of narcolepsy. To replicate whether incidence of NT1 in 2009-2011 were statistically increased compared to other years in the European population, we used the same data modeling approach as previously described in a Chinese study by Fang Han et al.^8^. Autoregressive integrated moving average (ARIMA) models were used on the data of 1995-2008 to forecast the numbers of NT1 in 2009-2011 with 95% predictive conference intervals (CIs). Then the ratios between the real and the predicted numbers of patients (i.e., *R=real-number/predictive-number*) and their 95% predictive CIs were calculated in 2009-2011, respectively. The incidence of NT1 was considered as *R*-fold significantly increased if the bottom of the 95% predictive CIs of *R* was larger than 1.

ARIMA models are suitable to fit the time series data and to forecast future data points in that series. However, ARIMA cannot use the data after the 2009 pH1N1 episode to predict the numbers in 2009-2011. We therefore used LOESS (locally estimated scatterplot smoothing methods)^18^, another model that allows us to exploit the entire dataset, both before and after 2009-2010 pH1N1 to predict the numbers of cases in 2009-2011. Similarly as the aforementioned analyses done with ARIMA models, we then predicted the numbers of NT1 onset in 2009-2011 using the LOESS models and calculated the ratios between the real and predicted numbers of patients and their 95% predictive CIs.

In addition, we divided the database into two subgroups, children and adolescent cases (age of starting EDS ≤18 yrs, n=256, f: 127, m: 129) and adult cases (age of starting EDS >18 yrs, n=252, f: 103, m: 149), and repeated the LOESS modeling in the two subgroups to further investigate whether the increased numbers of NT1 in 2009-2011 were age-specific. Since the delayed diagnosis is one of the major biases in the time series analyses as aforementioned, the ratios between the numbers of children and adult patients were calculated in each year. We used this artifice to find the genuine increase in either children or adult cases by cancelling out delayed diagnosis (i.e., we assumed that the delayed diagnosis equally influenced the numbers of children and adult patients in each year). We graphically analyzed the changes of the ratios from 1995 to 2016 and used box plots to depict outliers. The outliers could confirm whether the increased NT1 were age-specific in specific years after removing the delay bias.

The ARIMA models were built using the *R* package *forecast*^19^, in which the optimal models were selected automatically based on the bias-corrected Akaike information criterion (AICc)^20^. The LOESS models were 2-degree local polynomial regression and the model selections were done automatically based on AICc as well. They were built using the *R* package *fANCOVA*^21^. We used CIs rather than P-values to determine whether the prediction values of our models were significantly different from the real values in 2009-2011 (i.e., the results were significant if the 95% predictive CIs did not contain the real values), considering that P-values can only dichotomise significant or non-significant of hypothesis testing while CIs could inform both the range of predictions and the statistical significance^22^.

The data were expressed as means ± standard error (SE) unless indicated otherwise. Box plot was used in descriptive statistics to visually show the distribution of the data, including the median, interquartile range (IQR), the minimum (1^st^ quartile-1.5* IQR), the maximum (3^rd^ quartile + 1.5*IQR) and the outliers (data smaller than the minimum or larger than the maximum) of the data. All the analyses were done using *R* (version 3.5.3).

## Results

### Results of ARIMA models using data from 1995-2011

Combining all European countries, in total 39, 68 and 42 patients developed narcolepsy in 2009, 2010 and 2011, respectively. All tested patients (113 out of 149) showing EDS in 2009-2011 were HLA DQB1*06:02 positive. The 68 patients in 2010 was significantly 2.34-fold higher (95% CI: [1.79, 3.41]) than the 29 cases that were anticipated by the ARIMA prediction model (Fig.1).

**Fig.1.**
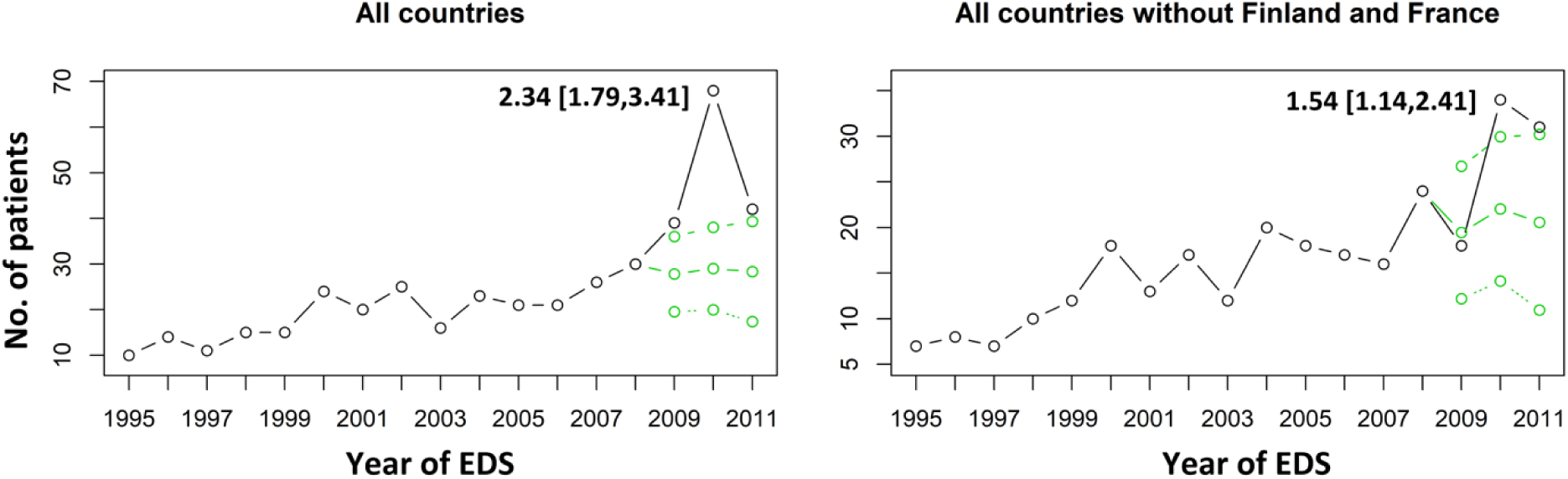
The predictions of ARIMA models combining all countries and in the countries without Finland and France. The predicted values and their 95% predictive CIs are marked as green circles, and the real values in black. The ratios and its 95% predictive CIs between the actual and the predicted values are shown in the figure.

Finland and France were two signaling countries previously reporting significantly increased NT1 IR in 2010 previously. The same ARIMA models were carried out in these two countries, respectively. The actual new cases in 2010 in Finland (15 patients) and France (19 patients) were significantly 9.78-fold (95% CI: [2.49, ∼]) and 4.07-fold (95% CI: [1.90, ∼]) increased compared to the predicted numbers, respectively. Considering that the significant result over all countries could be mainly driven by the strong effects of France and Finland, we repeated the ARIMA prediction model combining all countries except these two countries (Fig.1). The number of actual new cases remained significantly higher than the predicted number (1.54-fold higher, 95% CIs: [1.14-2.41]).

### Results of LOESS models using overall data from 1995-2016

The LOESS models included all data both before and after 2009-2010 pH1N1 to predict the numbers of new cases in 2009-2011 (Fig.2). The results confirmed the significant increases in patients in 2010: combining all the countries the increase was 2.54-fold higher (95% CI: [2.11, 3.19]) and after removing France and Finland the increase was 1.65-fold (95% CI: [1.36, 2.11]).

**Fig.2.**
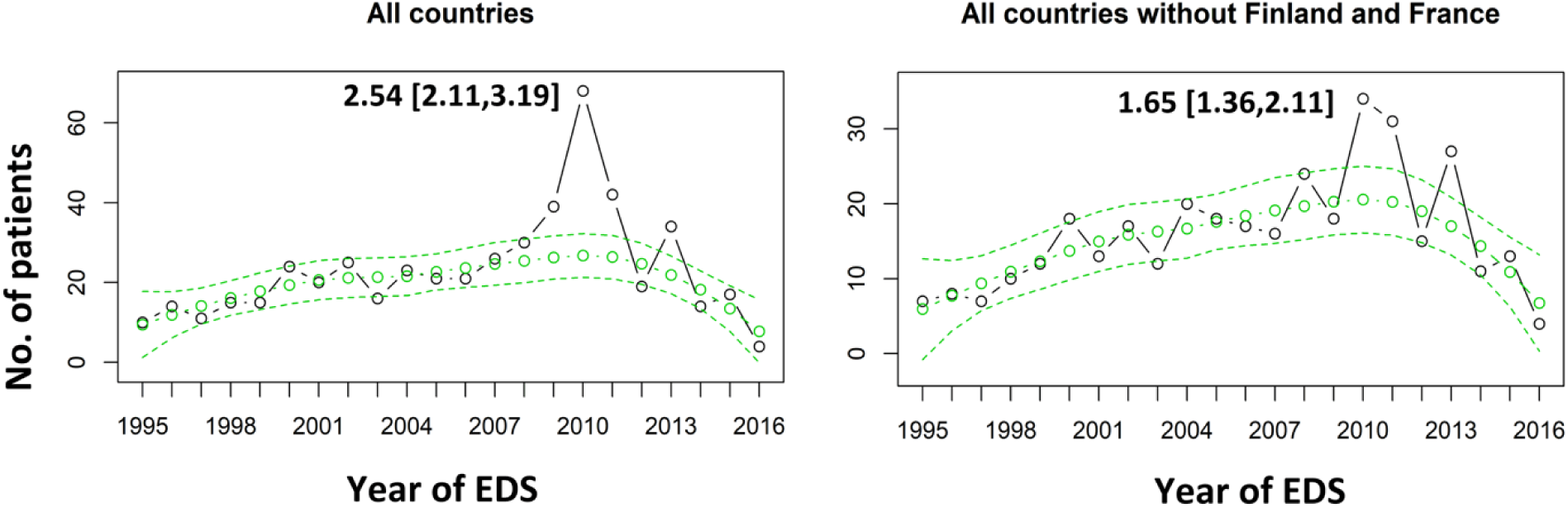
The predictions of LOESS models combining all countries and in the countries without Finland and France. The caption is the same as Fig.1.

The narrower predictive CIs of LOESS models compared to the ARIMA models (i.e., [2.11, 3.19] vs. [1.79, 3.41], [1.36, 2.11] vs. [1.14-2.41], as shown in Fig.2 and Fig.1) indicated that the overall LOESS models were indeed more robust than the ARIMA models in predicting the numbers of new cases because they also included the information after 2009 pH1N1.

### Results of LOESS models using overall data from 1995-2016 in individual countries

We further analyzed the data from each country individually using the more robust LOESS models. The results were shown in Fig.3. Significant increases were found in all the countries except for Italy and Switzerland in 2009-2011. The increases were 3.91-fold (95% CI: [2.75, 6.79]), 14.07-fold (95% CI: [7.19, 325.61]), 3.39-fold (95% CI: [1.75, 49.33]) and 2.03-fold (95% CI: [1.25, 5.43]) in 2010 in France, Finland, Spain and Czech Republic, respectively. Although significant increases in 2010 can be observed in the Netherland and Germany, the maximum increases were 1.92-fold (95% CI: [1.44, 2.89]) and 2.21-fold (95% CI: [1.47, 3.82]) in 2011 in these two countries, respectively.

**Fig.3.**
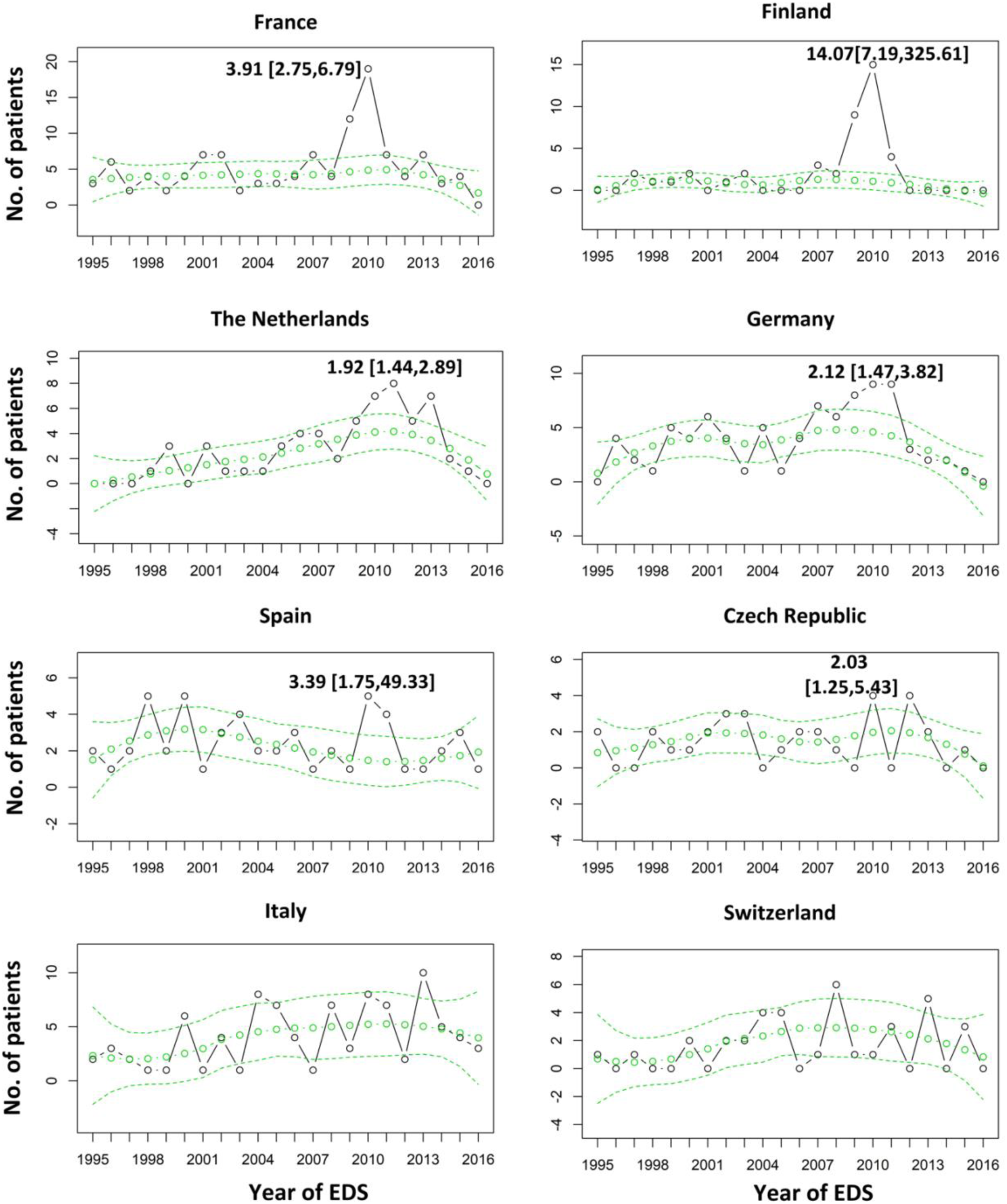
The predictions of LOESS models in each country. The caption is the same as Fig.1.

### Results of over-all LOESS models checking the age-specific increases in 2010 in all countries

The mean age of our children and adolescent patients was 12.08±0.24 yrs (IQR: 9.17-15.44 yrs). The mean age of our adult patients was 32.10±0.25 yrs (IQR: 24.23-38.44 yrs). Of children patients starting EDS in 2009-2010, 59.0% (46 out of 78) was diagnosed in 2009-2012. This proportion was 70.7% (46 out of 65) in adult patients (Fig. 4). Fig.4 also confirmed that some patients were diagnosed several years later after the 2009-2010 pH1N1, which is more evident in children/adolescents than in adults.”

**Fig.4.**
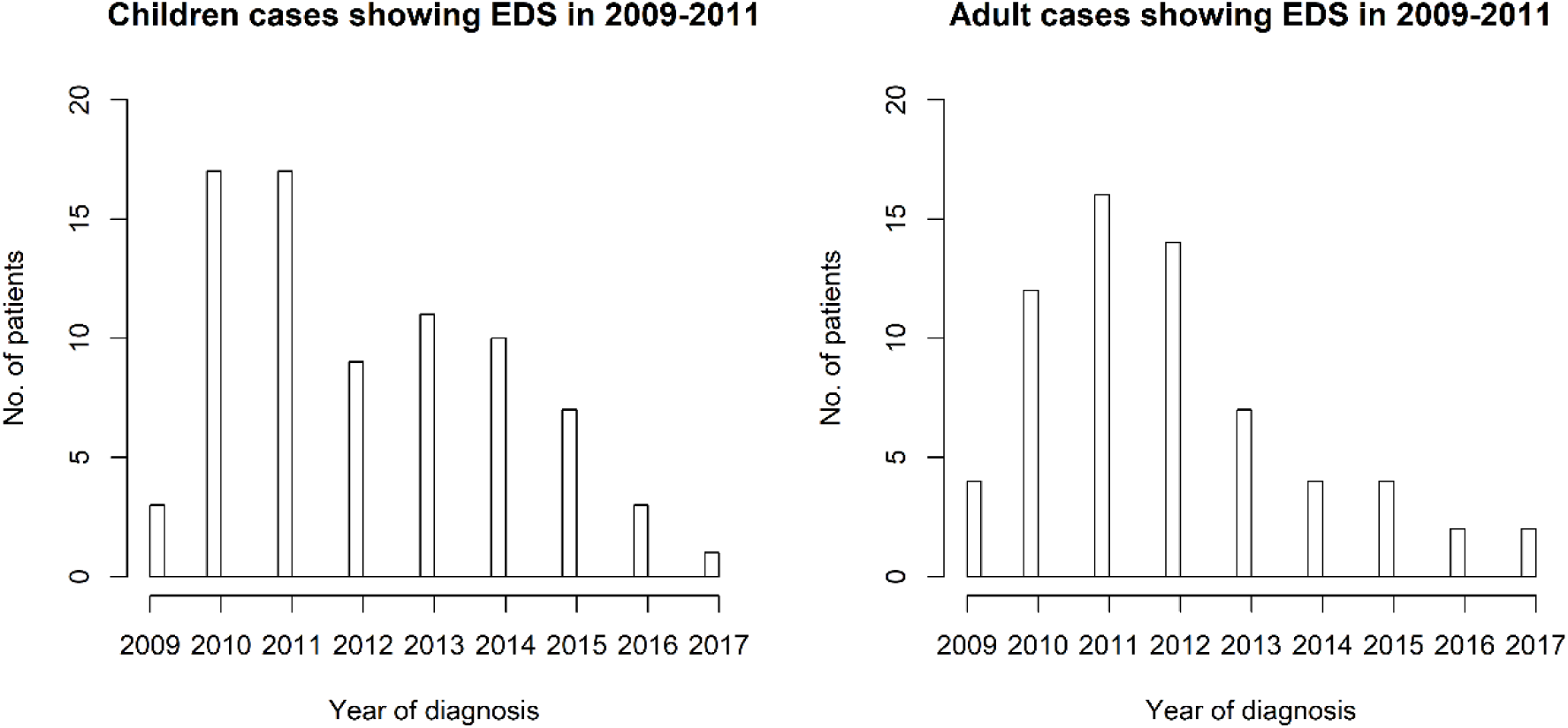
The year of diagnosis of children and adult patients starting EDS in 2009-2011.

Both in children/adolescents and in adults we found significant overall increases in the number of actual cases with EDS onset in 2010 (Fig.5) compared to the predicted numbers (2.75-fold, 95% CI: [1.95, 4.69] and 2.43–fold, 95% CI: [2.05, 2.97], respectively). It was remarkable to notice that specifically in children we could also find a 2.09-fold (95% CI: [1.52, 3.32]) increase in 2013, which was not shown in adults.

**Fig.5.**
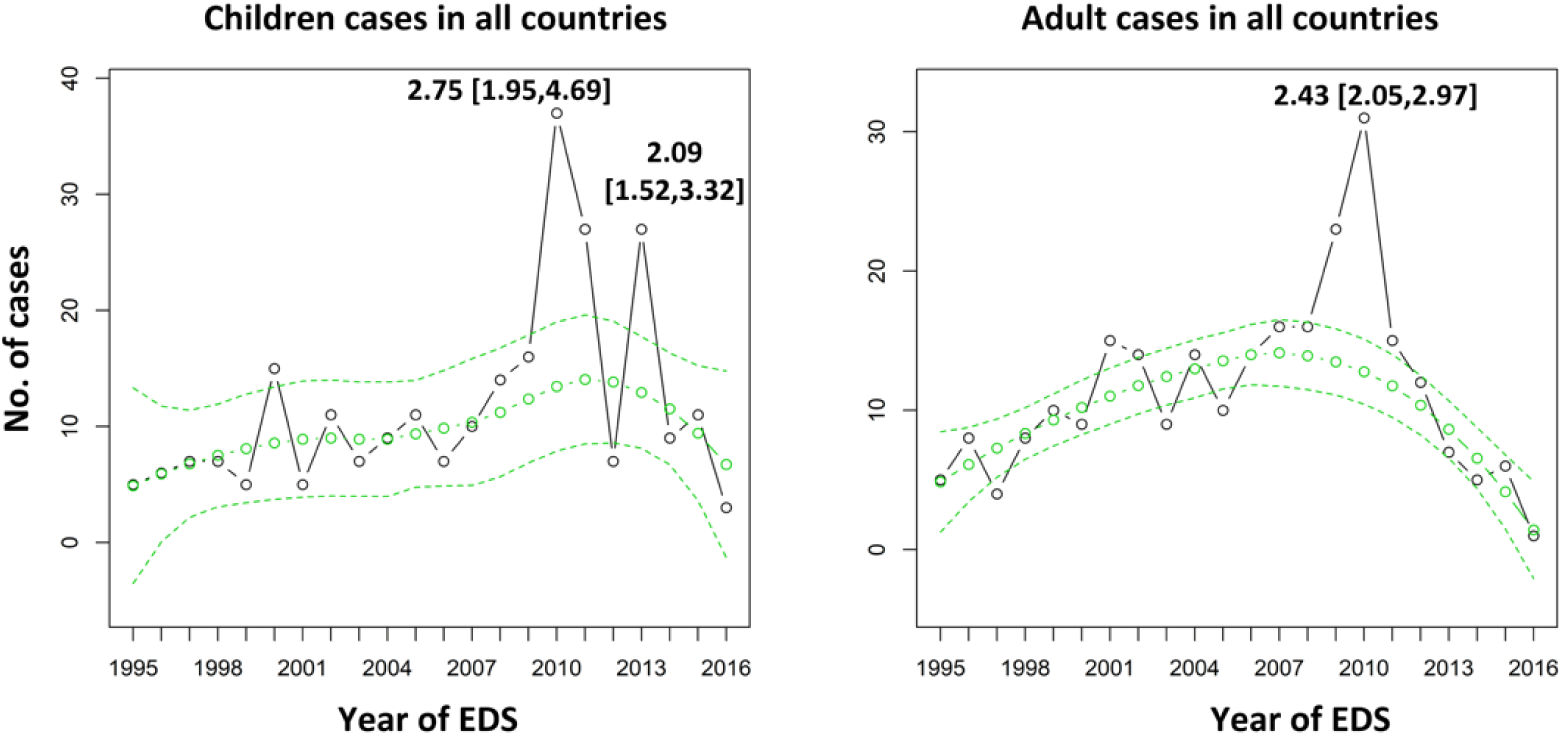
The predictions of LOESS models for children and adults cases in all countries. The caption is the same as Fig.1.

### Age-specific increases in NT1 in individual countries

The increase in 2013 in children and adolescent patients was unexpected. We further checked if the increases in 2010 or 2013 were age-specific for individual countries by modelling the numbers of children/adult patients using LOESS models (Fig.6), respectively. The results were:

**Fig.6.**
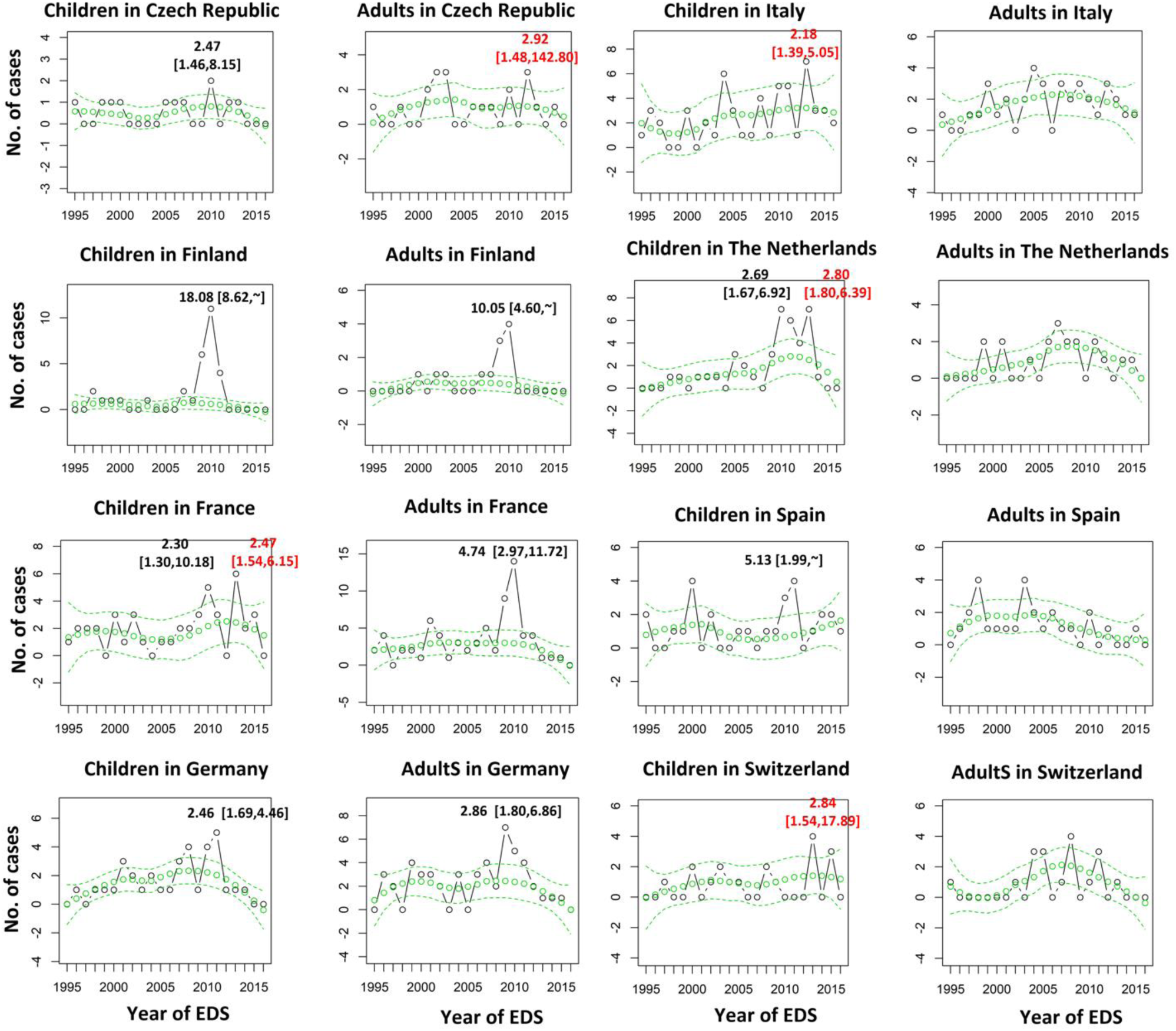
The predictions of LOESS models for children/adolescent and adult cases in each country. The predicted values and their 95% predictive CIs are marked as green circles, and the real values in black. The ratios and its 95% predictive CIs between the actual values and the predicted values are written in the figure, in black for the ratios in 2009-2011 seasons and in red for the ones in 2012-2013 season.

1. In 2009-2011 significant increases in the children and adolescent cases were found in Czech Republic (2.47-fold, 95% CI: [1.46, 8.15]), Finland (18.08-fold, 95% CI: [8.62, ∼]), France (2.30-fold, 95% CI: [1.30, 10.18]), Germany (2.46-fold, 95% CI: [1.69, 4.46]), the Netherlands (2.69-fold, 95% CI: [1.67, 6.92]) and Spain (5.13-fold, 95% CI: [1.99, ∼]). Significant increases in the adults cases in 2009-2011 were only found in Finland (10.05-fold, 95% CI: [4.60, ∼]), France (4.74-fold, 95% CI: [2.97, 11.72]) and Germany (2.86-fold, 95% CI: [1.80, 6.86]). Therefore the increases in Czech Republic, Spain and Netherlands in 2010 were age-specific for children/adolescent narcolepsy in 2010.
2. In 2009-2011 the maximum increases in children/adolescent and adults patients in Germany occurred in 2011 and 2009, respectively. In France and Finland the maximum increases in both children and adults patients were in 2010.
3. In 2013 significant increases in children/adolescent NT1 were found in Italy (2.18-fold, 95% CI: [1.39, 5.05]), the Netherlands (2.80-fold, 95% CI: [1.80, 6.39]), France (2.47-fold, 95% CI: [1.54, 6.15]) and Switzerland (2.84-fold, 95% CI: [1.54, 17.89]). Only in Czech Republic we found a significant increase (2.92-fold, 95% CI: [1.48, 142.80]) in adult NT1 in 2012. But the result should be interpreted cautiously considering the relative small number (n=3) of adult patients. Thus the increase in 2013 was age-specific.

### Age-specific increase in 2013 was confirmed by analyzing the children/adult ratios that was not biased by the delayed diagnosis

The changes of the ratios (i.e., children/adults) confirmed that the increase in 2013 was specific for childhood narcolepsy (Fig.7). The highest value was seen in 2013 (i.e., 27/7=3.86) recognizable as an outlier in the box plot (Fig.7). Fisher exact test showed that the ratio of children cases over adult cases was 4.15-fold higher in 2013 compared to in other years (P-value=0.0005, 95% CI: [1.72, 11.53]).

**Fig.7.**
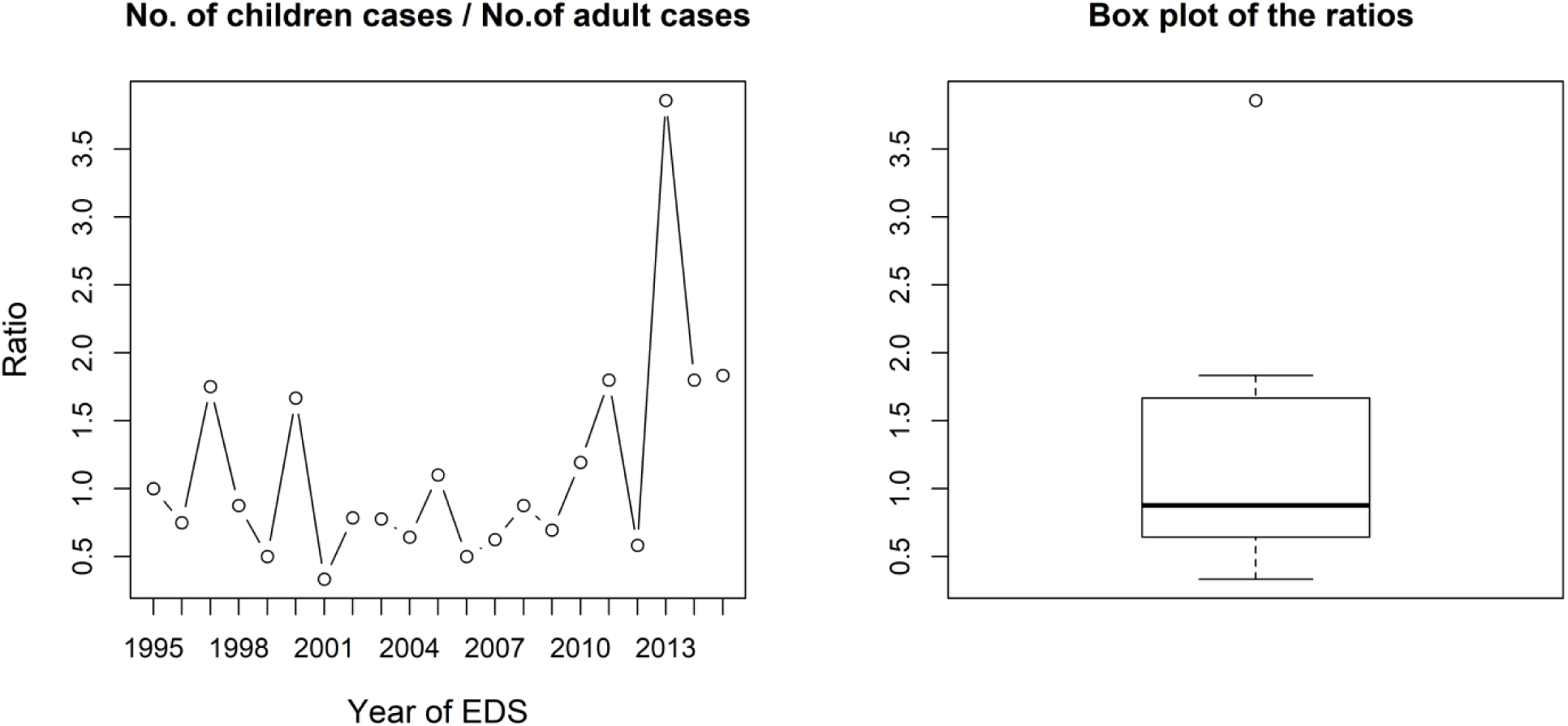
The changes of the ratios between children and adult patients from 1995 to 2015. The median of the ratios is 0.88 and the 1st and 3rd quartiles of the boxplot are 0.64 and 1.67, respectively.

All tested children/adolescent cases (19/27) showing EDS in 2013 were HLA DQB1*06:02 positive. The majority of these cases (16/25=64%, the other 2 patients had missing data of the cataplexy onset) developed cataplexy within 3-4 months after EDS in 2013, and this proportion was 70.6% (24/34, the other 3 patients had missing data of cataplexy onset) in 2010. 72.2% (13/18, the other 9 patients had missing data) of the children cases had EDS in April-June in 2013 (Fig 8). In 2010, 50% (17/34, the other 3 patients had missing data) of children patients showed EDS in January-March (Fig. 8). Taken the data from 2009-2010 together, 51% (24/47, the other 6 patients had missing data) patients had EDS in the 2009-2010 winter (i.e., December 2009 to March 2010) (Fig. 8).

**Fig.8.**
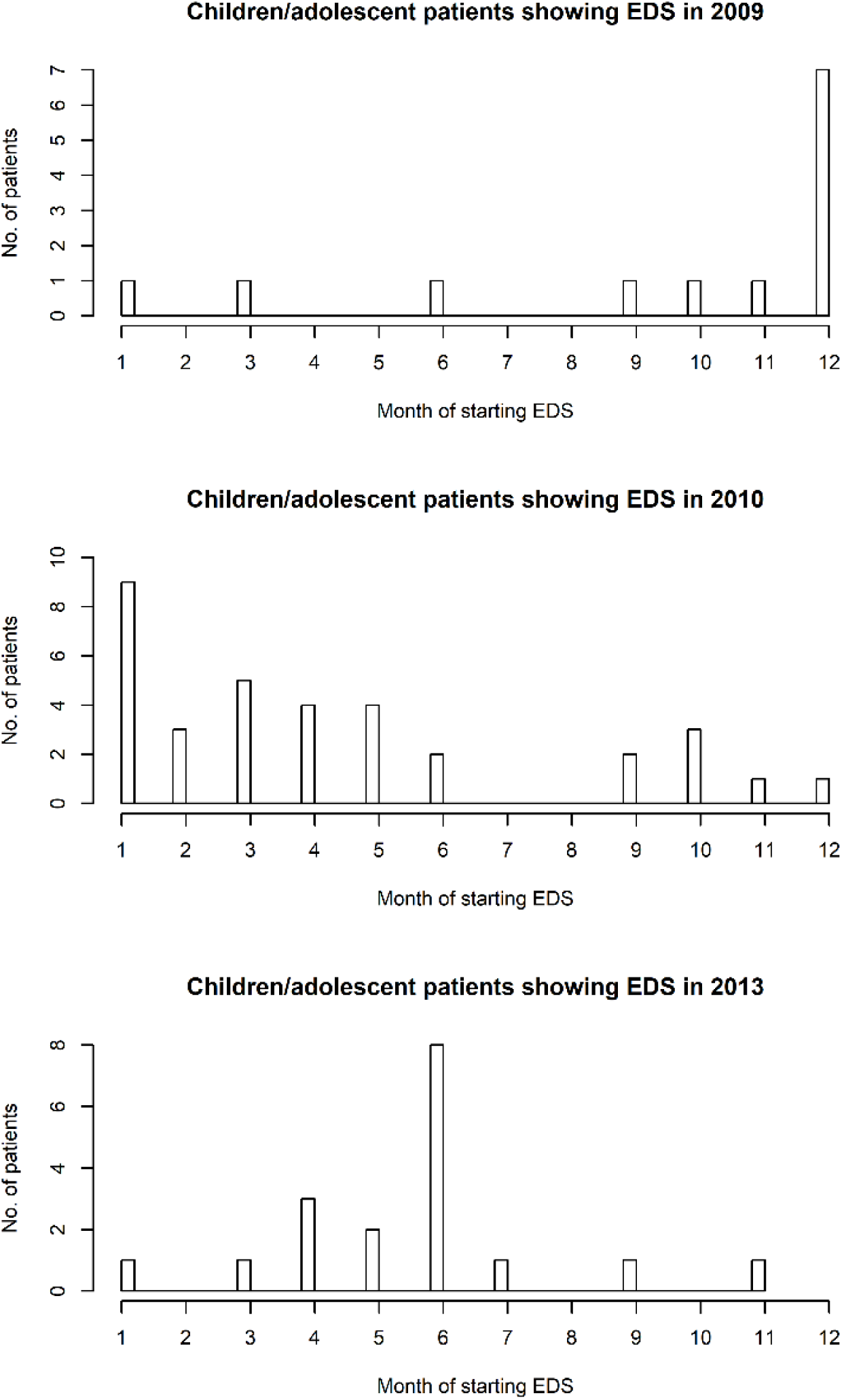
The seasonal number of children/adolescent cases in 2009-2010 and in 2013.

## Discussion

Using data-driven modelling approaches, we analyzed the changes in the number of the new NT1 cases (i.e., the onset of EDS) in the last two decades using the European Narcolepsy Network (EU-NN) database. As a major result we identified a new peak of NT1 incidence in 2013 that is age-specific for children. In addition we confirmed the peak in 2009-2010 pandemic H1N1 influenza that has already been identified in China and in some individual European countries. We consider our results as robust since we used a more sensitive model to analyze the data and, for the first time, took the delayed diagnosis into account, which was a major bias of previous studies. Several other aspects of our findings, in particular the age-specificity, the subacute disease onset of these de-novo children cases and the restrained use of Pandemrix after 2010 allow us to argue for a new epidemiological event triggering the increased NT1 cases in 2013.

### Methodological aspects: are our results robust

We applied two different models, first the ARIMA prediction models and second the more sensitive LOESS models. ARIMA models have already demonstrated an increased NT1 incidence in 2009-2010 H1N1 influenza season published in China^8^; Using exact the same prediction modelling we confirm an increased number of NT1 patients in 2010 Europe-wide. Unfortunately, the ARIMA models are less informative, because not all data can be included into the models. Due to inherent methodological reasons ARIMA models are powerful to forecast future data points based on previous data (“forward prediction”), whereas ARIMA is unable to use data following the forecasted data points (“backward prediction”) ^23^. In other words, we cannot include data after 2009-2011 to exploit their contributions to the prediction of the 2009-2011 incidences. The exclusion of data after 2009-2011 however is specifically problematic for diseases with delayed diagnosis like narcolepsy and is one of the major biases of previously published studies^7, 14^. Delays in diagnosing narcolepsy for months or even years makes it difficult to identify the exposures that contribute to disease development. We therefore run an additional, more sensitive overall prediction model (LOESS), since it incorporates all available data both before and after the 2009-2010 influenza pandemic. The longer follow-ups (from 2012 to 2016) in our database also allow us to identify more patients with a disease onset in 2009-2011. Accordingly, a considerable proportion (35.7%, 51 out of 143) of the patients starting EDS in 2009-2011, finally diagnosed after 2012 (see Fig.4), are included in our analysis. The superiority of the LOESS over ARIMA models is evident in the narrower predictive CIs. In summary our analysis using a more sensitive model with all available data (including the ones after 2010) provides us a better picture of the yearly incidences of NT1 in Europe.

### Increases of NT1 cases in 2010 and 2013

This study, for the first time, finds a significantly increased number of new patients in 2013. It also confirms the peak in 2010 that has been previously reported in Finland^5^ and France^4^. The 2013 increase is age-specific and specifically robust in France, the Netherlands, Italy and Switzerland. The significant increases in 2010 are seen in more countries such as The Netherlands, Germany, Spain and Czech Republic, some of these countries were not discovered in previous studies (i.e. The Netherlands and Spain^7, 14^). We also replicate the insignificances already reported in Italy^7^ and Switzerland^15^ in 2010, and find confirmative data supporting an age-specific temporal evolution of NT1 in children vs. adults as previously reported in Germany ^24^.

Some important differences between the increases in 2010 and 2013 should be explicitly mentioned:

1. The countries showing increased number of NT1 cases in 2010 and 2013 are not identical. Only France and Netherlands show an increase in both years, whereas in Italy and Switzerland it is just present in 2013.
2. The increased NT1 onset in 2013 is age-specific for children and show a typical subacute disease onset as previously described in immune-mediated narcolepsy. In 2010 the increase is found in both adults and children patients in most countries.
3. 50% of new patients in 2010 develop symptoms in winter (January-March), while in 2013 the onset mainly (72.2%) occurs in spring (April-June).

Both of our findings, the 2013 and the 2010 data provide several lines of arguments to further elucidate the potential association between narcolepsy and exposure to a vaccine or an infectious agent. The 2013 incidence peak strongly supports an epidemiological event in 2012-2013 triggering de-novo cases in childhood narcolepsy. The majority of these children cases (16/25=64%) developed cataplexy within 3-4 months after EDS consistent with the clinical descriptions of rapid symptom progression in immune-mediated narcolepsy^6^. This rapid evolution of our incident children cases makes it less likely that Pandemrix vaccination, which was no longer used after 2009-2010 pH1N1 according to the recommendation of European Medicines Agency, is also responsible for The NT1 increase in 2013. We can only speculate which type of epidemiological event may cause the 2013 peak. Recirculation of H1N1 or circulation of other/new influenza virus are options. Type B Influenza virus is one of the candidates as it more often impacts children^25, 26^, and its peak circulation is in late-February/early-March in 2013 in those countries for which we find the increased incidences^26-31^. The peak weeks of 2009-2010 pH1N1 circulation are in late November/early December in 2009 in Europe^32^. Both circulation peaks would fit the time lags of 3-4 months between influenza circulation and maximum increase of NT1 because the peak number of de-novo NT1 children is in winter in 2010 while it is in late spring in 2013. A new infection as trigger also fits the multiple hit hypothesis and would be compatible with a new peak in incidence in children soon after the 2009-2010 peak.

Two additional arguments derived from our 2010 data are in favor for a virus infection rather than Pandemrix vaccination for triggering narcolepsy in countries where Pandemrix was only used rarely (e.g. Germany) contrary to countries with high coverage of Pandemrix vaccination in 2009-2010 (e.g. Finland, Sweden and Ireland). In Germany the temporal evolution of narcolepsy is age-specific and different in children vs. adult cases. The maximum increase for children narcolepsy occurs in 2011 while it occurs in 2009 for adults (see Fig.6). Previous studies from Germany using a segmental linear regression analysis show an increased narcolepsy IR in children post-pandemic (maximum in 2011) compared to pre-pandemic^24^. Although the authors fail to find an overall increase in the IR in 2009-2011 in German adult cases, their data show that the maximum IR for adults is in 2009 and decreases after 2010. The overall vaccination coverage in Germany during 2009-2010 pH1N1 is estimated to be 8.1% and 7.8% in age groups older and younger than 14 years, respectively^33^. This low vaccination coverage together with our finding of the maximum increase occurring in 2009 rather than in 2010 in German adult cases suggest that H1N1 virus itself could be a triggering factor of narcolepsy. In two other countries, Finland and France (see Fig.6) the numbers of adult cases also start to significantly increase in 2009. Additionally, in the whole EU-NN database we could find that the number of adult patients in 2009 is already significantly increased compared to pre-pandemic, although the peak is in 2010 (Fig.5).

### Limitations and Perspective

We could not directly explore the pathophysiology of influenza/vaccination associated narcolepsy as the EU-NN database was not designed as a surveillance study and does not include the influenza and vaccination histories of the patients. We will further analyze in future studies, limiting to countries where vaccinations registries and individual vaccination histories are available. Second, for many reasons not all patients have been entered from all sleep centers in EU-NN database. We also lack information from some non-EU-NN member countries, such as Ireland, Norway and Sweden, where an association between Pandemrix and NT1 has been observed. Although we assume that our sample gives a representative figure about the European narcolepsy patients, a selection bias and influences by missing data are possible. Since the study is data-driven and not initiated by hypothesis it is reasonable to treat missing data as missing at random. By that they are less likely to bias the results and conclusions but of course we must be careful before making final inferences.

In spite of these limitations, our study still provides a novel approach, i.e., data-driven modelling, to investigate the potential triggers of narcolepsy. We find that the 2009-2010 pH1N1 may influence more European countries than we have known before. The unexpected increased children/adolescent narcolepsy in 2013 calls for more studies to further investigate the links between infectious agents (potentially, influenza type B Yamagata lineage), vaccination, genes and narcolepsy in 2013. The association of the population developing NT1 with the haplotype HLA DQB1*06:02 is one more argument in favor of the immune-mediated process involved in the pathophysiology of NT1 showing a possible connection between some active viral infections or attenuated forms of virus, on vaccines, and narcolepsy. In such context very cautious decision should be taken in account when vaccination in this HLA population group is considered. These studies will improve our knowledge of the pathophysiology of immune-mediated narcolepsy and the pathological links between vaccinations and narcolepsy.

## Data Availability

The datasets generated during and/or analysed during the current study are available from the corresponding author on reasonable request.

## Acknowledgment

The EU-NN database is financed by the EU-NN. The EU-NN has received financial support from UCB Pharma Brussels for developing the EU-NN database.

## Author Contributions

Study design: Zhongxng Zhang, Ramin Khatami, Jari Gool, Rolf Fronczek, Gert-Jan Lammers. Data analysis: Zhongxing Zhang. Paper writing: Zhongxing Zhang, Ramin Khatami, Jari Gool, Rolf Fronczek, Gert-Jan Lammers. Data acquisition: all the authors. All authors discussed the results and commented on the manuscript.

